# EPCs-derived conditioned medium mitigates chronic cerebral ischemic injury through the MIF-activated AKT pathway

**DOI:** 10.1101/2023.11.19.23298748

**Authors:** Ya-Wen Cheng, Ling-Yu Yang, Yi-Tzu Chen, Sheng-Che Chou, Kuo-Wei Chen, Yi-Hsing Chen, Chuan-Rou Deng, I-Chin Chen, Wan-Ju Chou, Chen-Chih Chang, Yong-Ren Chen, Hsiao-Lin Hwa, Kuo-Chuan Wang, Meng-Fai Kuo

## Abstract

**Background:** Chronic cerebral ischemia (CCI) is considered as a prelude to neurodegeneration. Endothelial progenitor cells (EPCs) have been implicated in revascularization and vascular repair in cerebral ischemic diseases. Due to the safety concern and the low survival rate of the transplanted cells, interest has shifted toward the paracrine effect of EPCs. Here, we investigate the effects of EPC-derived conditioned medium (EPC-CM) on the vascular and functional impairments in a rodent model of CCI and the mechanism via which the EPC-CM involves.

**Methods:** Bilateral internal carotid artery ligation (BICAL) was performed in rats to induce cerebral ischemia. EPC-CM was intracisternally injected 1 week after BICAL. The changes of the microvasculature and behavior were examined 3 weeks after BICAL. The EPC-CM was analyzed by cytokine array for the factors that involved in angiogenesis. The therapeutic effects and mechanism of the candidate factor was validated with oxygen-glucose deprivation-injured endothelial cells and EPCs *in vitro*.

**Results:** EPC-CM significantly improved the vascular, motor and cognitive impairments of the BICAL rats. Macrophage migration inhibitory factor (MIF) was identified as a key factor in EPC-CM involved in angiogenesis and anti-senescence. Furthermore, recombinant MIF protein mirrored the effects of EPC-CM on EPCs and ECs. These therapeutic effects were decreased by the co-treatment with EPC-CM and MIF-specific antibody both *in vivo* and *in vitro*. MIF operates through multiple pathways, including the AKT pathway, which plays a crucial role in cellular homeostasis. Inhibiting the AKT pathway diminished the protective effect of MIF in the CCI model.

**Conclusions:** We demonstrated that EPC-CM protected the chronic ischemic rat brain from ischemic injury and promoted functional recovery in rats through MIF-mediated AKT pathway, which indicated that EPC-CM may serve as an alternative potential therapy in chronic cerebral ischemia.

## Introduction

Chronic cerebral ischemia (CCI) results from inadequate blood flow to the brain. The decrease in cerebral perfusion induces brain tissue damage, cognitive impairment, and neurodegeneration, eventually causing ischemic stroke and Alzheimer’s disease^1-4^. Addressing this issue and finding ways to support neuron survival and neurological recovery is crucial due to the global impact of cerebral ischemic disease.

Endothelial progenitor cells (EPCs) are circulating cells capable of becoming endothelial cells for vascular regeneration. They play a vital role in repairing damaged blood vessels in ischemic disease^5-7^. EPCs secrete a wide range of growth factors, chemokines, cytokines, and extracellular vesicles that contribute to angiogenesis, reconstruct the vascular microenvironment^8-10^, and improved the recovery of neurological function after cerebral injury^11, 12^. Although cell-based therapy has shown promising effects in preclinical models after stroke, the administration of EPCs may have adverse side effects. Therefore, conditioned medium (CM) collected from EPCs has emerged as an alternative potential cell-free therapeutic agent^13-15^.

Our previous study demonstrated that the administration of EPCs combined with indirect revascularization protected against chronic cerebral ischemic injury^16^, but the therapeutic effects of EPC-secreted factors have not been investigated in preclinical models of CCI. We investigated the therapeutic effect of secreted factors from EPCs on CCI and the molecular mechanism involved in the neurorepair process. Many proteins and cytokines are secreted from EPCs, and we focused on macrophage migration inhibitory factor (MIF) among the secreted factors^17^. MIF is a pleiotropic protein that has the characteristics of a cytokine that is secreted by different types of cell, including neural stem/progenitor cells and EPCs^18, 19^. In addition to being recognized as a regulator of the immune response, increased studies have demonstrated that MIF is involved in a variety of signaling pathways important for the maintenance of cellular homeostasis, such as promoting cellular survival, angiogenesis, and anti-senescence^18, 20^.

In this study, we used a bilateral internal carotid artery ligation (BICAL) rat model as the CCI animal model and an *in vitro* cell-based system to determine whether EPC-CM promotes the recovery of brain tissue injury and vascular repair and further investigated the molecular mechanism involved in this process. The results demonstrated that EPC-CM can ameliorate ischemic injury and promote vascular repair in the brain via MIF-mediated AKT pathway activation.

## Methods

This study included the performance of animal experiments and obtaining EPCs from fresh human umbilical cord blood, and was approved by the Institutional Animal Care and Use Committee of National Taiwan University (Approval no. IACUC-20180430) and the Institutional Review Board of National Taiwan University Hospital (Approval no. IRB-201204074RIC and IRB-202012174RIND), Taipei, Taiwan. All surgical procedures were performed according to the Guide for the Care and Use of Laboratory Animals (issued by the National Institutes of Health) and approved by the Committee.

### Detailed information of the materials and methods

The detailed information of the materials and methods of the *in vivo* and *in vitro* studies were provided in the Supplementary Materials and Methods.

### Animal model of chronic cerebral ischemia: bilateral internal carotid artery ligation (BICAL)

Male Wistar rats (200–250 g, body weight) were randomly assigned to the BICAL surgery or treatment groups, and littermate control. The BICAL procedure was performed as described previously^16^.

### Treatment algorithm

The sham and BICAL animals received an injection of 40 μl vehicle, i.e, 0.9% normal saline into the cisterna magna, while the treatment groups received intracisternal injection of EPC-CM (40 μl) or EPC-CM+ MIF Ab (anti-MIF antibody 2 μg mixed with 40 μl EPC-CM) 24 hrs after BICAL procedure. The body temperature was maintained stable for 3 hrs after the treatment with vehicle, EPC-CM, or EPC-CM+ MIF Ab.

### Microcirculation assessment

Study and quantification of brain surface microcirculation 3 weeks after BICAL surgery was performed as described previously^16^.

### Microvasculature density

The microvascular densities were measured three weeks after BICAL surgery. The capillary microcirculation on the rat brain surface was evaluated using a CAM1 capillary anemometer (KK Technology, UK) with a high-resolution (752 x 582 pixels) monochrome charge-coupled device (CCD) video camera. The microvasculature density was calculated using the De Backer’s score^21^.

### Rotarod performance test

The rotarod test was performed as previously described^21^. Rats were pre-trained for three days at a speed of 4 rpm, 3 sessions per day for 5 min on the instrument (Panlab Rota Rod, Harvard Apparatus) before BICAL surgery. Three weeks after BICAL surgery, the rats were placed on the instrument and the latency to fall was measured under continuous acceleration (4 to 40 rpm for 600 secs) by observers blinded to group assignment.

### Novel Object Recognition (NOR) test

We conducted the NOR test to assess short-term recognition memory that originally described by Ennaceur and Delacour procedure^22^.

### Statistical analysis

Data collection and analysis were carried out by investigators who were blinded to the experimental conditions. Data are expressed as the mean ± standard deviation (SD). Data was analyzed for normality by the Shapiro–Wilk normality test. One-way analysis of variance (ANOVA) followed by a Post HocTest (Tukey’s multiple comparisons test) was used for normally distributed data. Bar graphs and statistical analysis were performed by GraphPad Prism 7.0 software (GraphPad Software, USA). A P-value of <0.05 was considered a statistically significant difference.

## Results

### EPC-CM treatment rescues the vascular impairment induced by BICAL

The microcirculation on the cortical surface, including the artery (a) and vein (v), showed significantly sparse in BICAL rats as compared with control rats. In addition, arterioles (a) on the brain surface exhibited markedly diffuse vasoconstriction in BICAL rats, resulting in ischemia (Figure 1A, middle panel). Notably, impairment of microvasculature and arterioles were rescued by treatment with EPC-CM (Figure 1A, right panel). As microvascular networks supply oxygen to brain tissue, we detected the microvasculature density, regional blood flow, and partial pressure of brain tissue oxygen (PtO_2_) in BICAL rats treated with/without EPC-CM. These measures, which had notably declined in the BICAL rats, were restored with EPC-CM treatment, indicating an improvement in cerebral perfusion (Figure 1B-D). Taken together, these data suggested that EPC-CM rescues the vascular impairment induced by BICAL.

**Fig.1.**
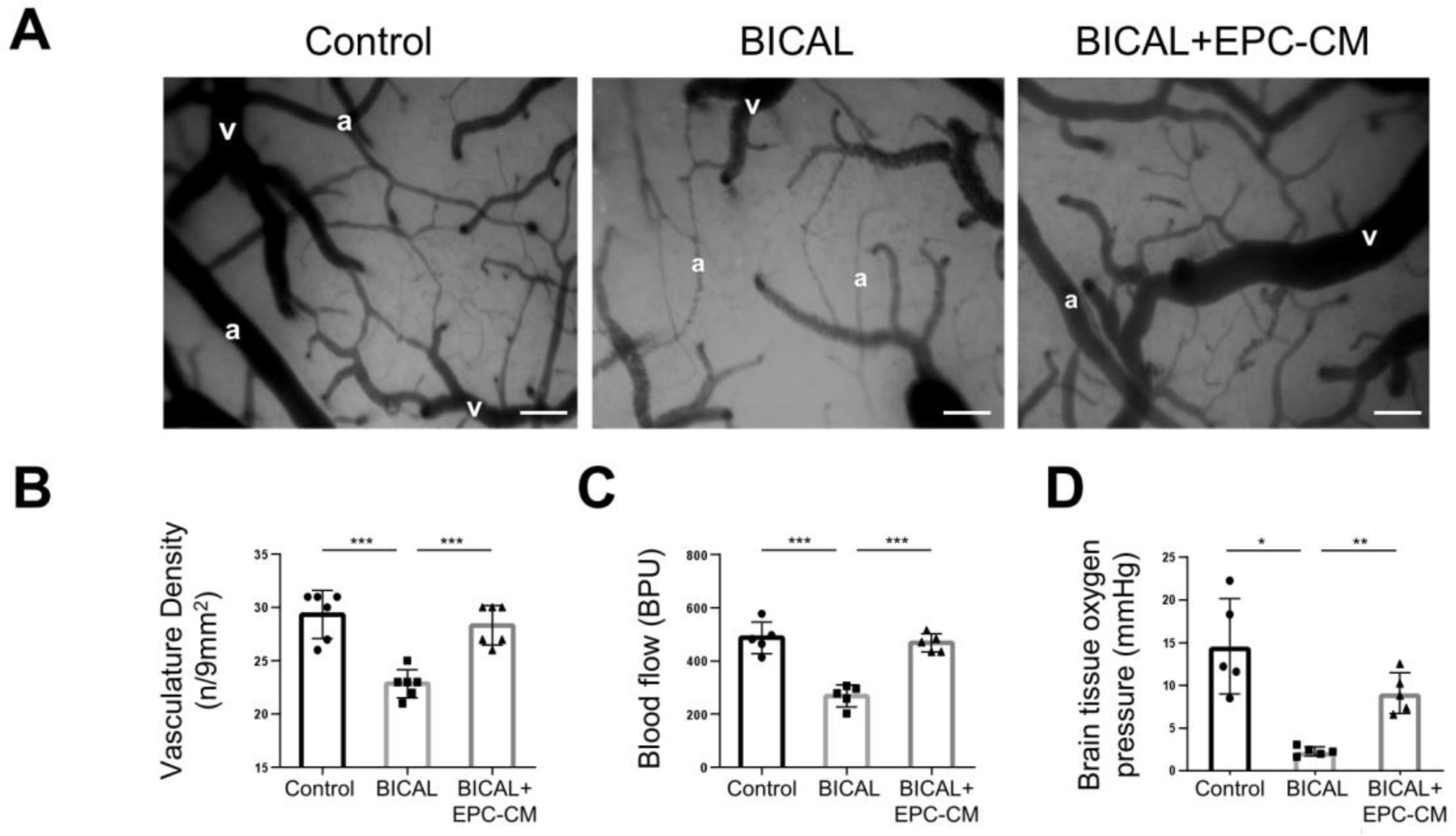
EPC-CM treatment rescues the vascular impairment induced in BICAL rat. **A,** The artery (a) and vein (v) were observed on the brain surface by the videoscopic view. The microcirculation on the cortical surface was significantly decreased in BICAL-rat. Scale bar = 50 μm. **B,** The effects of EPC-CM after BICAL on the cerebral microvasculature density was quantified by using a CAM1 capillary anemometer. **C** and **D**, The effects of EPC-CM on the microvascular system after BICAL were evaluated by regional blood flow (**C**) and the partial pressure of brain tissue oxygen (PbtO_2_) (**D**). Data was showed as the mean ± SD, and analyzed using by One-way ANOVA. * *p* < 0.05, ** *p* < 0.01, *** *p* < 0.001, n = 5–6 in each experiment.

### EPC-CM and MIF protein promote angiogenesis in OGD-treated endothelial cells (ECs)

To investigate the critical factor in EPC-CM that is responsible for mediating vascular and neural repairs in BICAL rats, cytokine array of EPC-CM was performed that showed macrophage migration inhibitory factor (MIF) among the secreted factors of EPC-CM as a putative candidate for promoting angiogenesis and cell survival^23, 24^ (Figure 2A, Table S1). Therefore, we tested the role of MIF in ECs by using human umbilical vein endothelial cells (HUVECs) treated with/without EPC-CM or MIF recombinant protein (rMIF) subjected to oxygen-glucose deprivation (OGD) to mimic chronic ischemic conditions. We investigated cell survival in HUVECs treated with OGD for 24 hrs and found no significant differences between each group, indicating that the condition did not cause significant cell death (Figure 2B). Next, we validated the angiogenesis function of the ECs by performing tube formation and migration assays. As shown in Figure 2C and 2D, compared to normoxia, OGD treatment significantly inhibited tube formation and cell migration. Interestingly, OGD cells treated with EPC-CM or rMIF showed similar increases in tube formation and cell migration (Figure 2C-2D). The data indicated that both EPC-CM and rMIF increase angiogenesis in the *in vitro* chronic ischemic model.

**Fig2.**
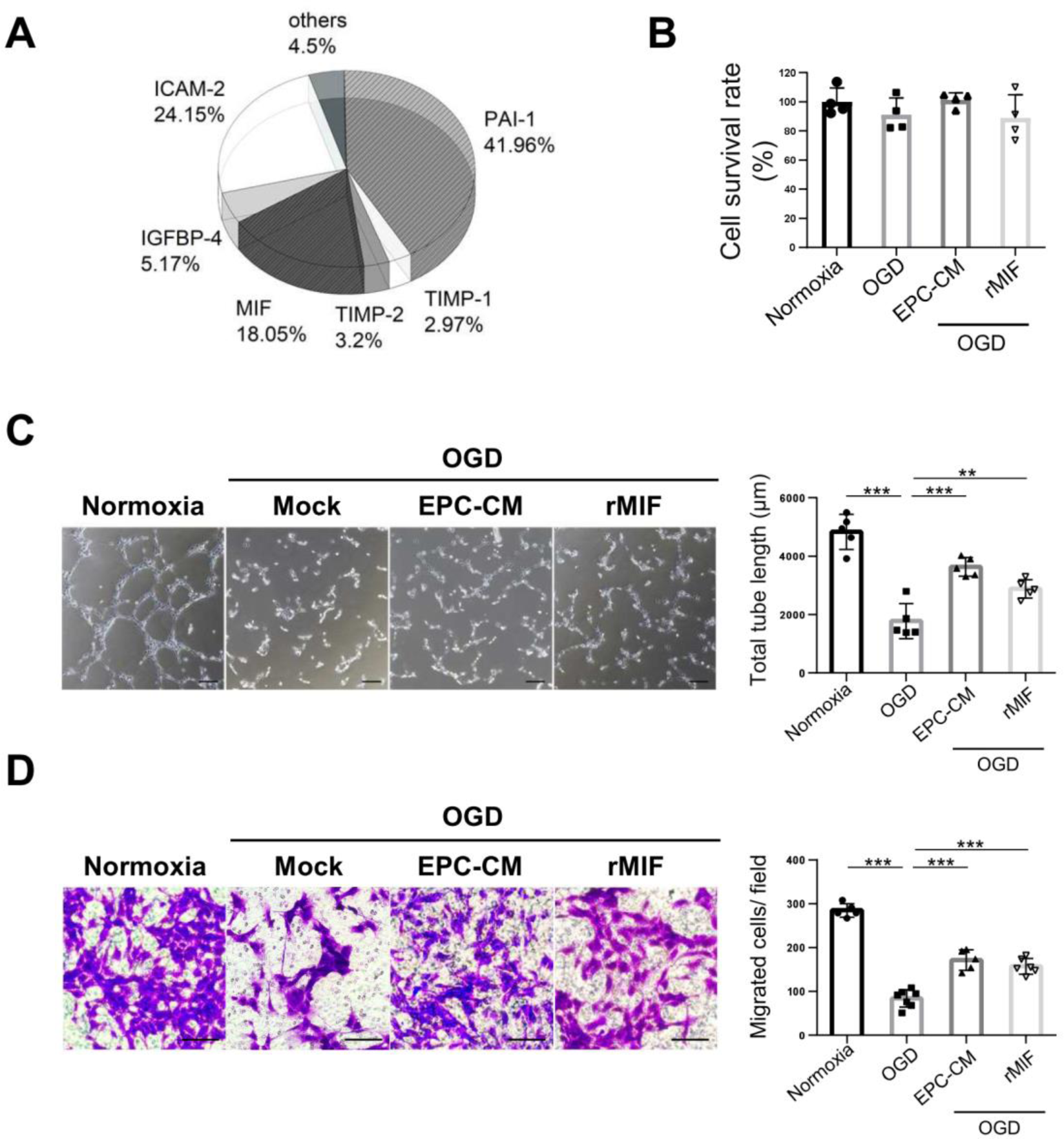
Both EPC-CM and MIF protein promote the angiogenesis ability in OGD-treated endothelial cells. **A,** Cytokine profile in the EPC-CM was determined using Quantitative Cytokine Quantibody Human Array 4000 (RayBiotech, Norcross, GA). **B,** The cell survival ratio of the OGD-treated HUVEC cells with/without EPC-CM or recombinant MIF treatment was detected by CCK-8 assay at OD 450nm and normalized with normoxia group. **C,** Representative images exhibited the tube formation in HUVEC cells with different treatments (left panel), and the average total length of tubes was quantitatively analyzed by 5 fields randomly (right panel). **D,** Representative images of the crystal violet staining of the HUVECs passed through the transwell membrane (left panel). The migrated cell numbers were quantitatively analyzed by 6 fields randomly (right panel). (C), (D) Scale bar = 100 μm. Data was showed as the mean ± SD, and analyzed using by One-way ANOVA ** *p* < 0.01, *** *p* < 0.001.

### EPC-CM and MIF protein promote anti-senescence ability in ECs and EPCs *in vitro*

Senescence is associated with neurodegeneration diseases^25^. Senescent EPCs showed decreased proliferation and angiogenesis. Therefore, we determined the anti-senescence effect mediated by EPC-CM or rMIF on EPCs and ECs. Young EPCs or ECs were treated with prolonged hydrogen peroxide (H_2_O_2_) to induce senescence, followed by treatment with EPC-CM (20%) or rMIF (100 ng/ml). As shown in Figure 3, H_2_O_2_ treatment significantly increased the number of senescent cells compared with that in the control (Figure 3, lane 1 vs. lane 2 in the bar graphs). Notably, EPC-CM or rMIF treatment reversed the senescence level in EPCs (Figure 3A) and ECs (Figure 3B). These data indicated that EPC-CM or MIF may promote angiogenesis by rejuvenating senescent EPCs or ECs after ischemic damage.

**Fig 3.**
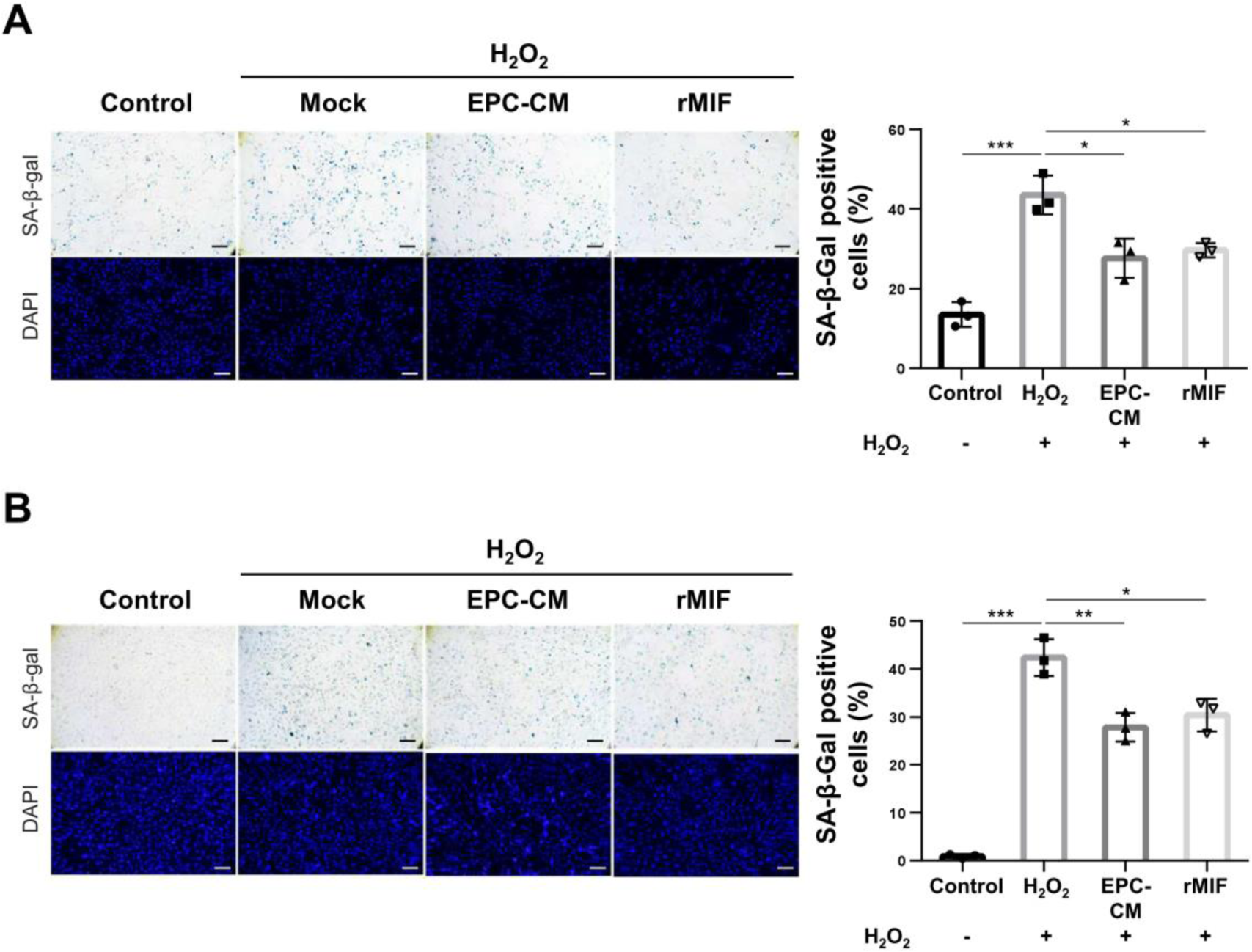
Both EPC-CM and MIF protein protect against the H_2_O_2_-induced senescence of ECs and EPCs. **A and B,** Representative bright micrographs of SA-β-gal staining of H_2_O_2_ -treated cells. Senescence of the young EPCs (**A**) or the HUVECs (**B**) (passage <10) was induced by H_2_O_2_ treatment, followed with EPC-CM (20%) or recombinant MIF protein (100 ng/ml) treatment (left panel). The senescence ratio was quantitatively analyzed of the SA-β-gal positive cells normalized with DAPI staining. The data was comparted with H_2_O_2_ group (right panel). Scale bar = 100 μm. Data was showed as the mean ± SD, and analyzed using by One-way ANOVA. * *p* < 0.05, ** *p* < 0.01, *** *p* < 0.001.

### Transduction of EPCs with lenti-hMIF increases cell angiogenesis and anti-senescence *in vitro*

To further confirm the role of MIF in angiogenesis and anti-senescence, we generated the human MIF-expressing EPCs by lentivirus, and validated the hMIF expression by Western blotting (Figure S1A). Angiogenesis and the anti-senescence capacity were tested in the hMIF-expressing EPCs compared with the lentiviral vector-transduced EPCs (Mock). As shown in Figure S1B and S1C, hMIF-expressing EPCs rescued tube formation and migration with OGD treatment as compared with OGD-treated cells infected with lentiviral vector. In addition, hMIF-expressing EPCs also restored the senescence level after H_2_O_2_ induction (Figure S1D), consistent with Figure 3. These data proved that MIF is an important protein that regulates angiogenesis and anti-senescence in EPCs.

### MIF is an important factor in EPC-CM that promotes angiogenesis and brain functional recovery in chronic ischemic events

To validate the role of MIF in the recovery of neurological function regulated by EPC-CM, we used anti-MIF antibody (Ab) to specifically block the effect of MIF in EPC-CM. While EPC-CM treatment significantly restored the tube formation (Figure 4A) and migration capacity (Figure 4B) inhibited by OGD treatment (Figure 4, lane 2 vs. lane 3 in the bar charts), the effect of EPC-CM was reduced by MIF Ab treatment (Figure 4, lane 3 vs. lane 4 in the bar charts). The data indicated that MIF is the key factor in EPC-CM that promotes angiogenesis and anti-senescence capacity.

**Fig 4.**
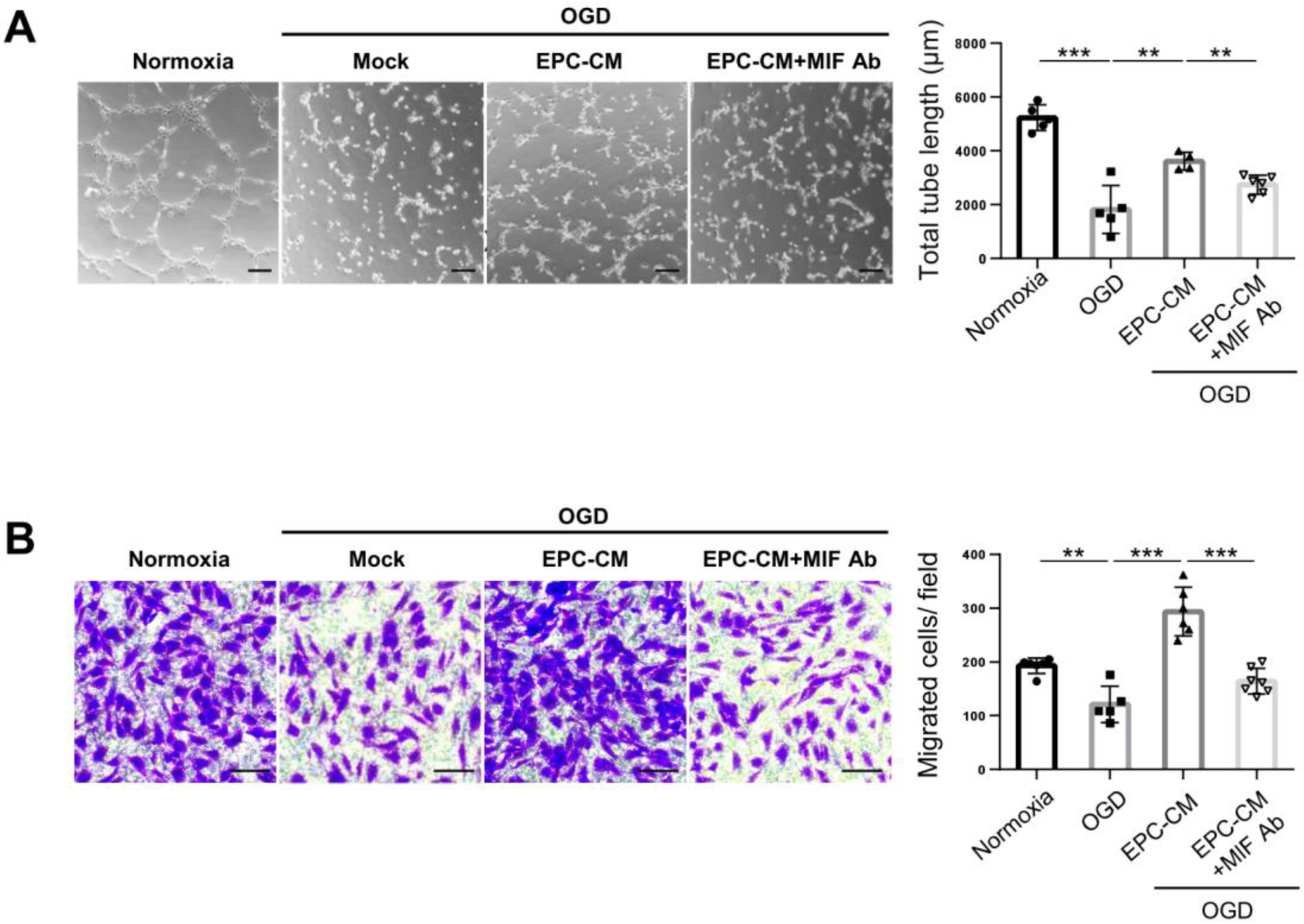
Neutralization of MIF repressed the angiogenesis mediated by EPC-CM treatment in vitro. **A,** Representative images of tube formation was examined by Matrigel assay (left panel). The total tube length in five random fields was quantified by MetaMorph software and showed as bar graph (right panel). Scale bar = 100 μm. **B,** Determination of the HUVECs passed through the transwell membrane stained with the crystal violet (left panel). The migrated cell numbers were quantitative analyzed by 6 fields randomly (right panel). Scale bar = 100 μm. Data was showed as the mean ± SD, and analyzed using by One-way ANOVA. ** *p* < 0.01, *** *p* < 0.001.

To further study the effect of MIF in the CCI animal model. Wistar rats were randomly separated into four groups (control, BICAL, BICAL+EPC-CM, BICAL+EPC-CM+MIF Ab) and underwent BICAL surgery followed by EPC-CM or EPC-CM+MIF Ab treatment as previously described (Figure 5A). The blood flow and PtO_2_ were examined for the repair of the microcirculation. According to Figure 5B and 5C, EPC-CM showed a great vascular repair effect on blood flow and PtO_2_, which was significantly repressed by MIF-Ab co-treatment, indicating that MIF plays an important role in EPC-CM-mediated vascular repair. Furthermore, we investigated the effects on the motor and cognition functions of BICAL rats treated with/without EPC-CM or EPC-CM+MIF Ab with rotarod test and the novel object recognition (NOR) assay, respectively. EPC-CM treatment reduced both motor coordination deficits and recognition memory impairment after BICAL, which were abolished by MIF Ab co-treatment (Figure 5D-E). These data indicated that EPC-CM is a potential cell-free therapy for CCI and MIF is the key factor in EPC-CM that promotes vascular repair.

**Fig 5.**
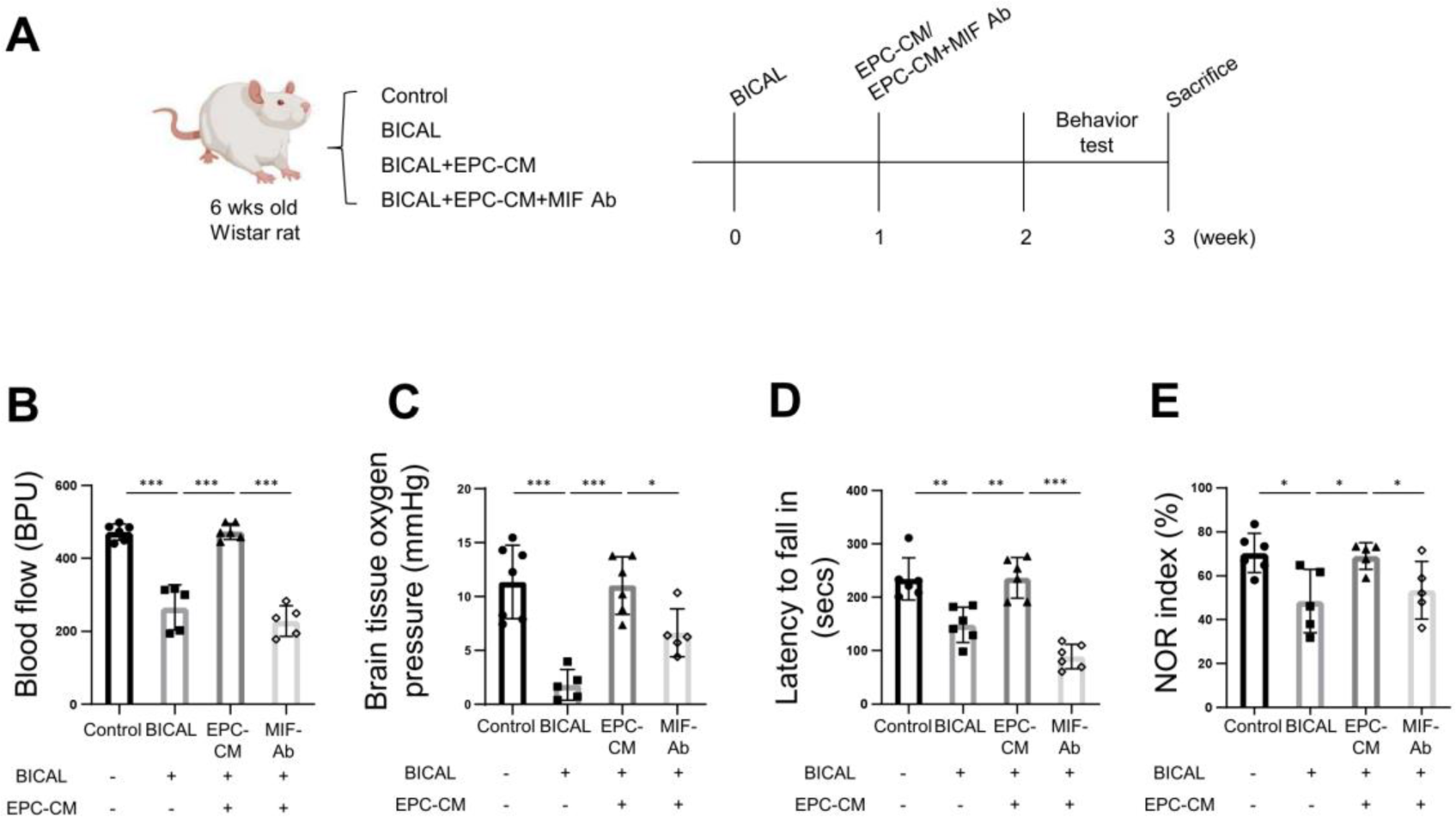
MIF is the key factor in the EPC-CM to promote vascular repair and protects the motor function and cognition after BICAL. **A,** The schematic representation of the experimental process. **B** and **C,** Determination of the vascular function of BICAL-rat treated with EPC-CM or EPC-CM+MIF Ab by analyzed the regional blood flow (B) or the partial pressure of brain tissue oxygen (PbtO_2_) (C) (n=5 in each group). **D,** The effects of EPC-CM and EPC-CM+MIF Ab on the motor function was evaluated by rotarod test (n=6). **E,** The recognition memory function in BICAL rat treated with/without EPC-CM or the EPC-CM+MIF Ab was detected by NOR test. The duration of time spent exploring the novel object was normalized with total time spent exploring both objects (n>5). Data was showed as the mean ± SD, and analyzed using by One-way ANOVA. * *p* < 0.05, ** *p* < 0.01, *** *p* < 0.001.

### MIF promotes angiogenesis and anti-senescence by activating the AKT pathway

Having established the role of MIF in promoting angiogenesis and anti-senescence by *in vitro* system, we investigated the underlying mechanism. Previous studies demonstrated that MIF increased angiogenesis and anti-senescence by mediating the phosphatidylinositol 3’ -kinase (PI3K) /AKT signaling pathway and mitogen-activated protein kinase (MAPK)^23, 26^. Therefore, we investigated the regulation of MIF or EPC-CM on ERK1/2 and AKT activity in EPCs. EPCs treated with rMIF or EPC-CM both showed significant increases in ERK1/2 and AKT phosphorylation, consistent with previous studies (Figure 6A). Next, we detected ERK1/2 and AKT activity with/without EPC-CM or rMIF treatment under OGD conditions. We found that EPC-CM or rMIF treatment increased the AKT activity inhibited by OGD in ECs and EPCs but not the ERK1/2 pathway (Figure 6B). The data indicated that AKT is the critical pathway regulated by MIF to promote angiogenesis and anti-senescence. To validate this hypothesis, we used a PI3K/AKT inhibitor, LY294002, to inhibit AKT activity, and the inhibitory efficacy was confirmed by Western blotting (Figure 6B, lane 5). As shown in Figure 6C and 6D, the tube formation and cell migration promoted by rMIF treatment was abolished by AKT inhibitor co-treatment. In addition, the anti-senescence effect mediated by rMIF was repressed by the AKT inhibitor (Figure 6E). Taken together, these data suggested that activation of AKT plays an important role in MIF-mediated neurologic repair by increasing angiogenesis and anti-senescence in CCI.

**Fig 6.**
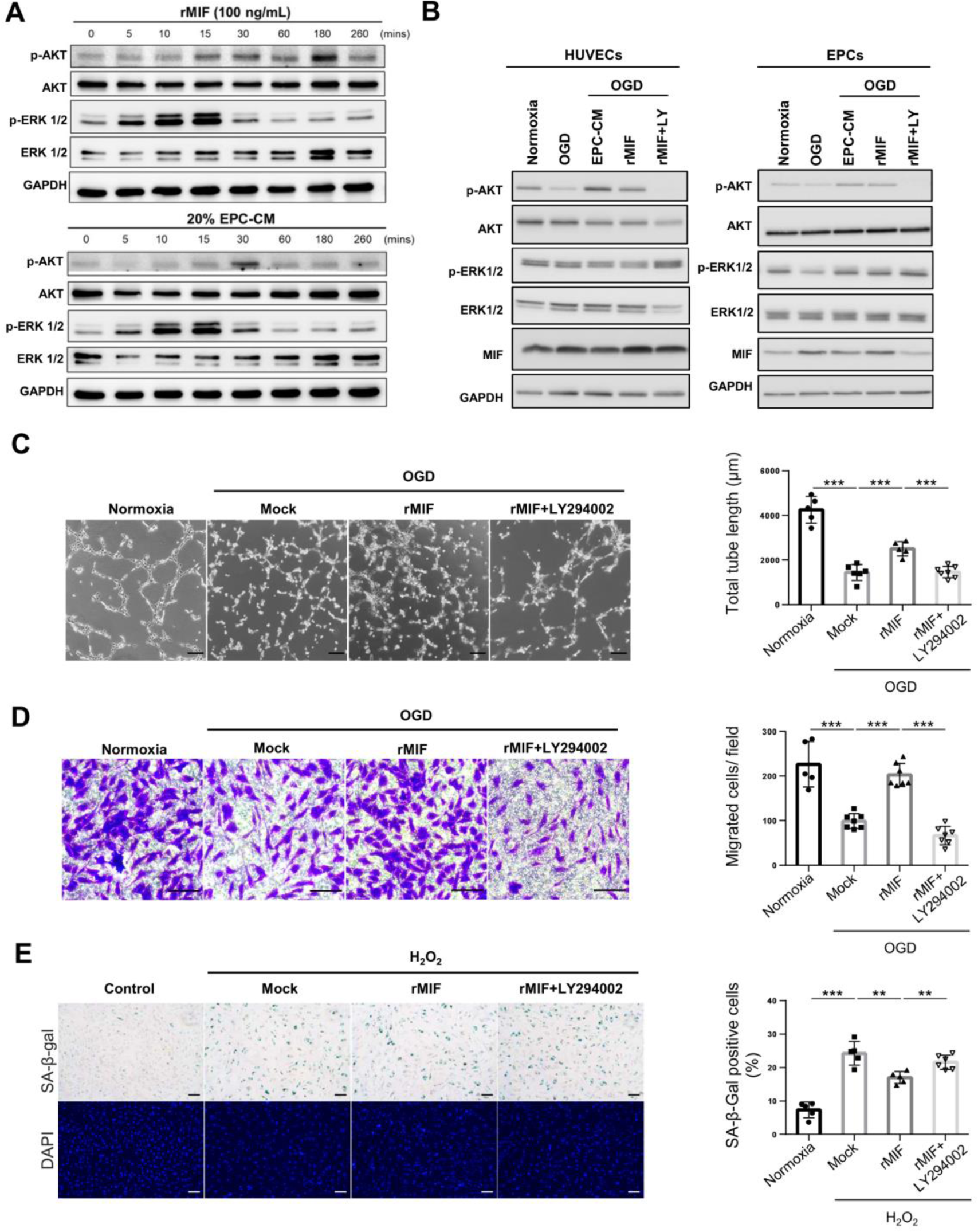
MIF promotes the angiogenesis and anti-senescence via activating the AKT pathway. **A,** Determination the AKT and ERK1/2 activity upon the treatment of EPC-CM or recombinant MIF protein in different time points in the EPCs. The EPCs were treated with rMIF (100 ng/ml) or EPC-CM (20%) for the indicated time and harvested the cell lysates for Western blotting analysis. **B,** Detection of the AKT and ERK1/2 activity in OGD-treated cells incubated with EPC-CM, or rMIF, or rMIF+LY294002. HUVECs (left panel) or EPCs (right panel) were incubated with/without EPC-CM (20%), or rMIF (100ng/ml), or the rMIF+LY294002 (5nM) at 0.5 % O_2_ incubator for 24 hrs. The cell lysates were collected to detect the AKT and ERK1/2 activity by using the Western analysis. **C,** Representative images of the tube formation of the OGD-treated cells incubated with different chemicals (left panel). The average total tube length of five random fields was quantified and showed as bar graph (right panel). Scale bar = 100 μm. **D,** Representative images of the HUVECs passed through the transwell membrane stained with crystal violet (left panel). The migrated cell numbers were quantitative analyzed by 6 random fields (right panel). Scale bar = 100 μm. **E,** Images of the SA-β-gal staining of H_2_O_2_-induced senescent cells treated with different chemicals in HUVEC cells. The senescence ratio was the SA-β-gal positive cells normalized with the DAPI in 6 independent fields. The data wad comparted with the H_2_O_2_ group (right panel). Scale bar = 100 μm, Data was showed as the mean ± SD, and analyzed using by One-way ANOVA. ** *p* < 0.01, *** *p* < 0.001.

## Discussion

Chronic cerebral ischemia (CCI) is highly correlated with neurodegenerative diseases and increases the mortality and disability rate in aging patients. Unfortunately, effective therapies for CCI are limited. In this study, we showed that conditioned medium derived from EPCs enhanced microcirculation, and cognitive and motor functions of CCI animals. We further identified MIF, a pleiotropic cytokine, as a critical factor in EPC-CM that exhibits angiogenesis and anti-senescence activities, facilitating tissue recovery in both *in vivo* and *in vitro* stusies. Mechanistically, EPC-CM promoted angiogenesis and anti-senescence in EPCs and ECs through MIF-mediated AKT activation. Overall, we demonstrated the therapeutic actions of EPC-CM in vascular and brain functional improvement in experimental CCI model.

Vascular repair is critical for cerebral protection after ischemic insults. EPCs, as the precursor cells of ECs, play a pivotal role in repairing injured vessels and participate in endothelial regeneration^27, 28^. Despite their attractiveness as a therapeutic approach for ischemic diseases, challenges such as immunogenicity, tumorigenic potential, and formation of emboli limit the clinical application of EPC transplantation^29^. The detailed mechanism by which EPCs contribute to vascular repair is still unclear. There are two mechanisms by which EPCs promote vascular repair. One is that EPCs migrate to the ischemic region and differentiate into ECs to repair damaged vessels. The other is that the recruited EPCs secrete protective paracrine factors to promote vascular repair^30^. The latter mechanism, particularly relevant for its stability, safety, and low immunogenicity, has gained prominence. Evidence supports that EPC-mediated vascular repair via paracrine signaling, activating resident ECs, is more critical than direct differentiation into damaged vessels^13, 31, 32^. In current study, we demonstrated that the administration of EPC-CM facilitates vascular repair after BICAL. Furthermore, by analyzing the paracrine factors of EPCs with a cytokine array, we focused on the role of MIF in EPC-CM-mediated promotion of angiogenesis and cell survival after BICAL.

We used the BICAL model, a CCI animal model, to investigate the therapeutic effects of EPC-CM. The cerebral ischemia induced by BICAL was validated by microcirculation changes, including decrease of vascular density, regional blood flow, and PbtO_2_. The microcirculation changes were significantly ameliorated by the administration of EPC-CM. Notably, EPC-CM treatment also rescued the impaired motor and recognition functions in BICAL rats that suggested the role of EPC-CM in neuronal protection after ischemic insults. The EPC-CM cytokine profile highlighted MIF as a key factor in promoting vascular and neuronal repairs. Intriguingly, MIF-specific antibody treatment inhibited the therapeutic effects mediated by EPC-CM, reinforcing the pivotal role of MIF in vascular repair and neuronal recovery.

MIF was first identified as an inflammatory cytokine and is now recognized as a protective factor that involves in diverse physiological processes, including angiogenesis, antioxidant activity, and cell survival^19, 33^. Many studies have demonstrated that MIF participates in the recovery of ischemic injuries^34-37^. MIF can suppress oxidative stress by its redox activity or inhibit the transcription of P53 to decrease cell apoptosis and senescence. MIF also promotes angiogenesis^38-40^. However, other studies have reported that MIF aggravates ischemic damage^41, 42^. Inácio *et al*. found a smaller infarct size 7 days after stroke in MIF-knockout mice^42^. These studies pointed out that MIF has different roles during ischemia, either promoting neuronal recovery by inhibiting cell death or being harmful to neurons by increasing the inflammatory response in ischemic region, and the mechanism underlying the different roles of MIF is still unclear. One possibility is that higher expression of MIF may activate broad signaling pathways and inflammation. In the rodent permanent middle cerebral artery occlusion (MCAO) models, MIF showed detrimental effects, which increased the permeability of blood brain barrier and the infarction volume with 3.3 µg/kg administration in Liu’s study^43^, whereas 0.9 ng MIF injected into mice via the ventricle (final 120 ng/ml concentration) promoted neurological recovery^34^. The MIF concentration in EPC-CM was 12 ng/ml, and treatment with 40 µl EPC-CM in BICAL rats showed significant protective effects in our study, which is consistent with the low dose of MIF administered in the ischemic stroke model^34^.

Furthermore, we showed the therapeutic effects mediated by MIF by increasing angiogenesis and anti-senescence in EPCs and ECs in the *in vitro* CCI model. Secreted MIF activates multiple cellular signaling pathways by binding to the cellular receptor CD74^44^. The interaction of MIF with CD74, triggers various signaling pathways, including the MAPK and PI3K/AKT pathways, which are crucial for angiogenesis and anti-senescence^23, 45^. In addition, MIF can bind to other receptors, CXCR2 and CXCR4, to regulate cell proliferation, survival, angiogenesis, and chemotaxis^46, 47^. Here, we demonstrated that MIF promotes angiogenesis via activation of the PI3K/AKT pathway to increase tube formation, cell migration, and anti-senescence.

Cerebral small vessel impairment is related to various diseases, especially cognitive dysfunction, vascular dementia, and stroke in elderly patients^48^. This study demonstrated the therapeutic effect of EPC-CM in animal model of CCI. Notably, in prior research, CM was administered for multiple times^9, 11^, while here, EPC-CM was delivered once, 7 days following BICAL surgery, yet still exhibited a substantial effect. This indicates that the EPC-CM used in our study had long-term effects on EPC-mediated reendothelialization and neovascularization, which might be due to EPC chemotaxis to the ischemic region mediated by MIF^16, 49^. An intriguing avenue for exploration is whether the vascular repair facilitated by EPC-CM could extend to various ischemic diseases. Further investigation is needed to elucidate the detailed mechanism of EPC-CM and MIF after ischemic events.

## Conclusion

In conclusion, our study in an animal model highlighted the therapeutic potential of EPC-CM for CCI. The pivotal role of MIF in angiogenesis and anti-senescence through the activation of the PI3K/AKT pathway provides mechanistic insights into the neuroprotective effects of EPC-CM. These findings hold promise for developing novel therapies for cerebral ischemic diseases.

## Data Availability

The data that support the findings of this study are available from the corresponding authors upon reasonable request.

## Data Availability

Yes, we had a statement regarding the availability of all data referred to in the manuscript

## Non-standard Abbreviations and Acronyms

CCI: chronic cerebral ischemia
BICAL: bilateral internal carotid artery ligation
EPC-CM: endothelial progenitor cell-derived conditioned medium
MIF: macrophage migration inhibitory factor
OGD: oxygen-glucose deprivation
ECs: Endothelial cells
HUVEC: human umbilical vein endothelial cell
SA-β-gal: senescence associated β-galactosidase
DAPI: 4’,6-diamidino-2-phenylindole
NOR: novel object recognition

## Acknowledgments

We thank the Animal Core Facility, Department of Medical Research, the Second and the Third Core Labs, Department of Medical Research, National Taiwan University Hospital for technical support during the study. YWC and KCW designed research, data interpretation, writing and revising of the manuscript; YWC performed *in vitro* experiments, acquired and analysis of the data; LYY, CRD, SCC, KWC, and YHC performed animal experiments. ICC and WJC contributed to Western blotting. YTC, YRC, HLH and CCC contributed to EPCs preparation. KCW and MFK contributed to overall direction, funding acquisition and critical revision of the manuscript.

